# Application of the Logistic Model to the COVID-19 Pandemic in South Africa and the United States: Correlations and Predictions

**DOI:** 10.1101/2022.01.12.22269193

**Authors:** David H. Roberts

## Abstract

We apply the logistic model to the four waves of COVID-19 taking place in South Africa over the period 3 January 2020 through 14 January 2022. We show that this model provides an excellent fit to the time history of three of the four waves. We then derive a theoretical correlation between the growth rate of each wave and its duration, and demonstrate that it is well obeyed by the South African data.

We then turn to the data for the United States. As shown by Roberts (2020a, 2020b), the logistic model provides only a marginal fit to the early data. Here we break the data into six “waves,” and treat each one separately. Five of the six can be analyzed, and we present full results. We then ask if these data provide a way to predict the length of the ongoing Omicron wave in the US (commonly called “wave 4,” but the sixth wave as we have broken the data up). Comparison of these data to those from South Africa, and internal evaluation of the US data, suggest that this current wave will peak about 18 January 2022, and will be substantially over by about 11 February 2022. The total number of infected persons by the time that the Omicron wave is completely over is projected be between 22 and 24 million.

## 1. Introduction

Since the spring of 2020 a pandemic of infection of a novel coronavirus (SARS-CoV-2) has overspread the world. Epidemiologists have struggled to describe adequately this pandemic and to predict its future course. This is a complex undertaking, involving the mathematics of epidemiology and a huge amount of uncertain data that serve as input to the models. In this paper we examine the ability of a simple epidemiological model, the *logistic model*, to describe the course of the pandemic in the Republic of South Africa and the United States. After showing that it is adequate to the task for South Africa, we derive a theoretical correlation between the growth rate of each wave of the pandemic and its duration, and show that it is well obeyed for the South Africa data. Finally, we discuss the use of this correlation to predict the course of the fourth (Omicron driven) wave in the United States, and show that it should die out very soon.

The data are from the World Health Organization (WHO 2022).

## 2. The Logistic Model

### 2.1. Derivation

The exposition in this section is from Roberts (2020a).

A simple model for the evolution of a pandemic is based on the logistic differential equation. This describes a pandemic that begins with a small number *f*_0_ of infected individuals, and subsequently spreads through a population. The motivation for this model is as follows. If the population of infected individuals as a function of time is *f* (*t*), simple exponential growth with growth rate *r* is determined by the differential equation

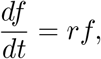

with the growing exponential solution

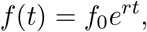

where *f*_0_= *f* (0). This is what happens with an infinite pool of subjects. However, for a finite pool of subjects, as the population of infected individuals grows the number of subjects available to be infected gets smaller. This is taken into account by modifying the exponential differential equation to become

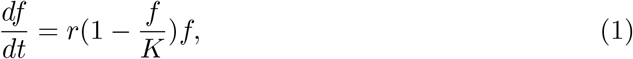

where *K* is the total available pool of individuals. The solution of this equation^2^ is

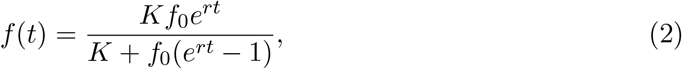

which satisfies the required limits *f* (0) = *f*_0_ and *f* (∞) = *K*. The time course described by Equation 2 is the familiar “S curve” used to describe bacterial growth and other phenomena (see Figure 3).

An analysis of the total number of cases as a function of time *f* (*t*) is just one way to compare model and data. Instead we can examine the number of new cases per day as a function of time; this is the time derivative of *f* (*t*).^3^ This is easily found by substituting Equation 2 into Equation 1,

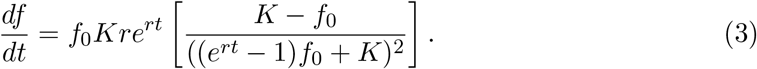

In either case, for the logistic model the three parameters to be adjusted are *f*_0_, *K*, and *r*. For waves with a substantial ongoing number of daily infections coming in, the model is augmented by adding a baseline level *N*_0_ that is also solved for from the data,

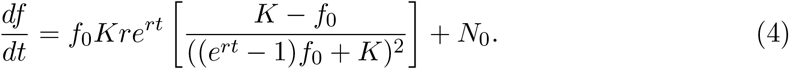

### 2.2. COVID data for the United States and the Republic of South Africa

The plots in this section show the total and daily distributions of the number of cases of COVID-19 in the United States and in South Africa (WHO 2022). Day 1 is 03 January 2020.

**Fig. 1.**
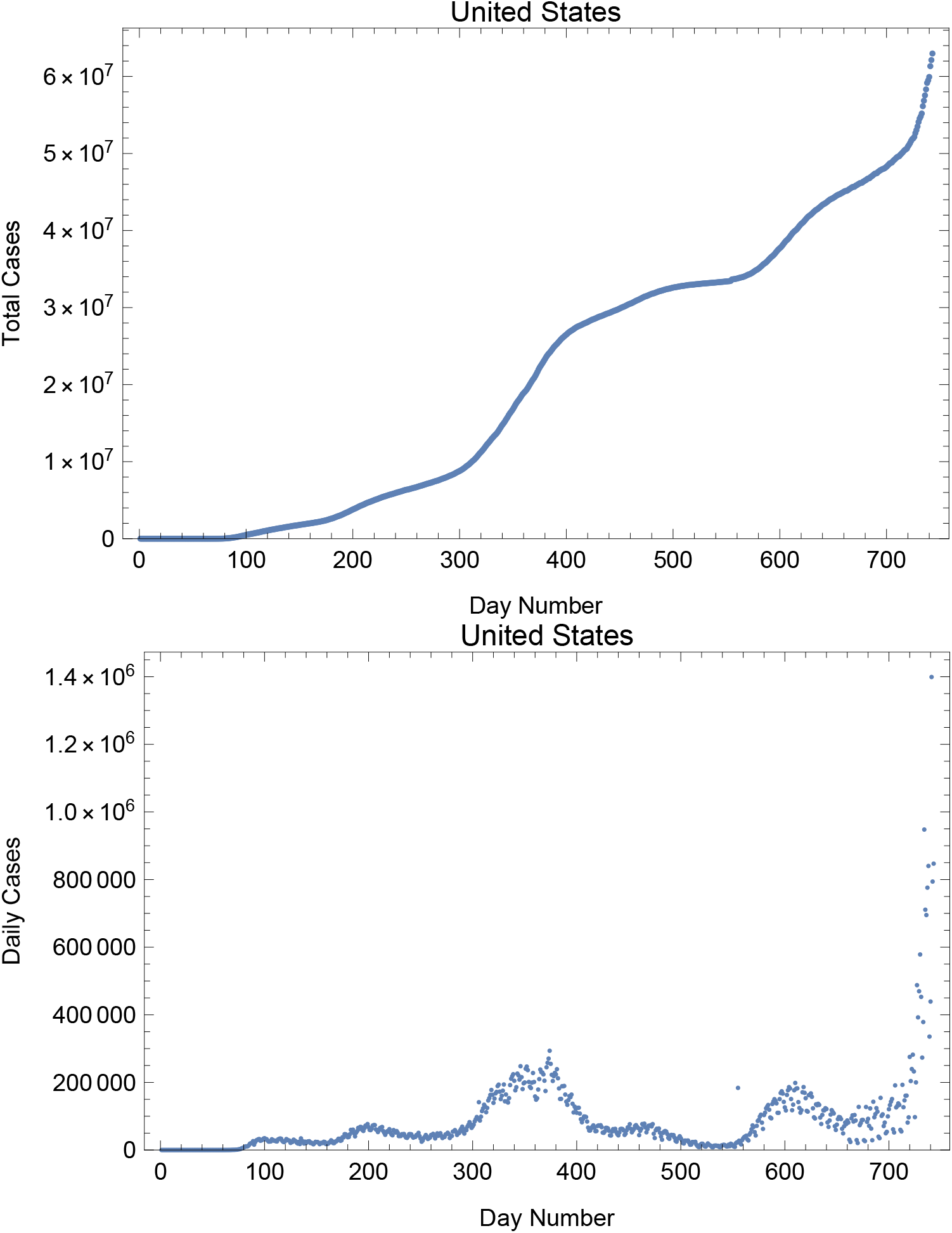
Cases data for the entire pandemic for the United States.

**Fig. 2.**
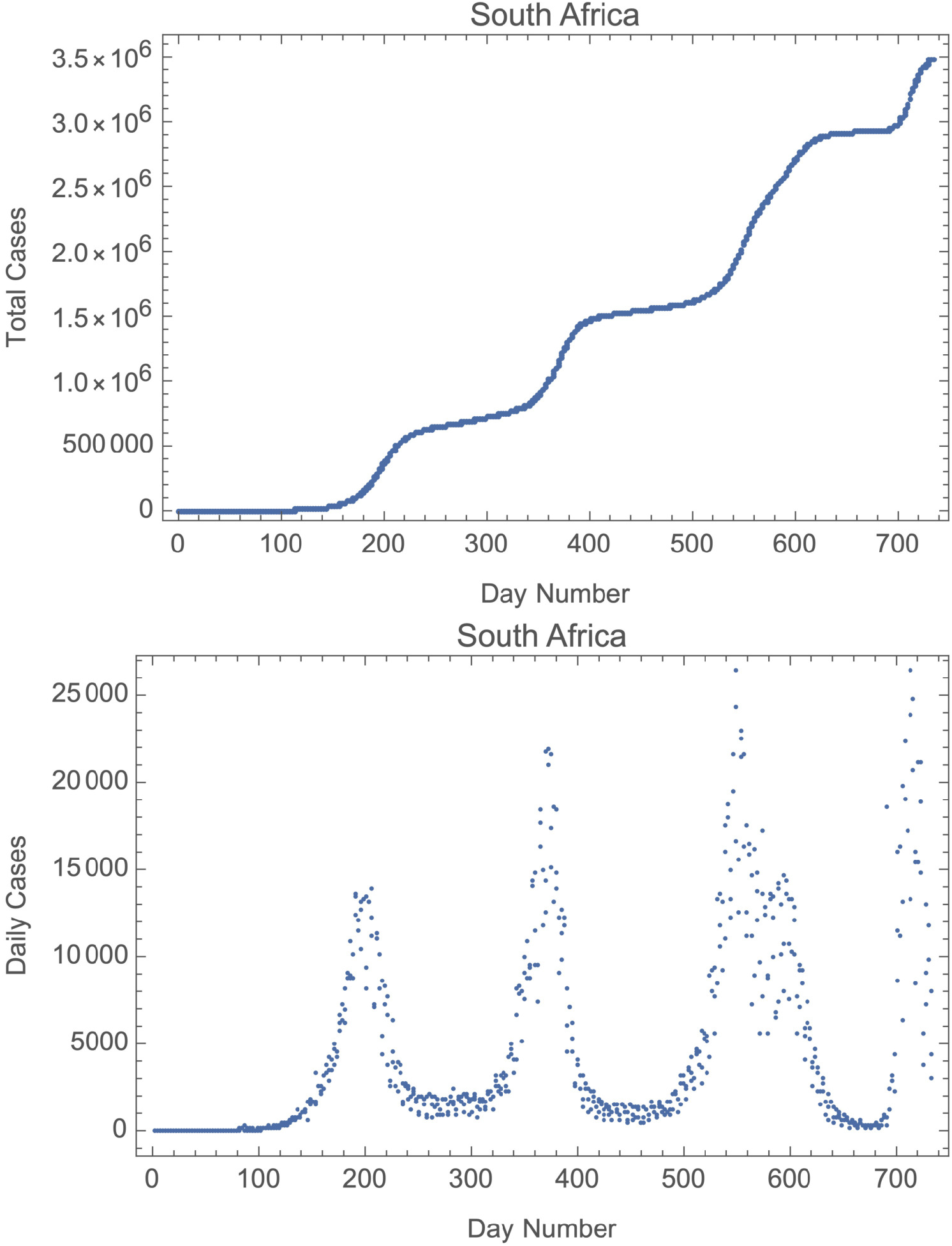
Cases data for the entire pandemic for the Republic of South Africa..

### 2.3. Fit to the COVID-19 Pandemic in the Republic of South Africa

When applied to the ongoing COVID-19 pandemic in the United States the logistic model does only a fair job of accounting for the actual history of the total number of infected individuals as a function of time in the first wave of COVID-19, which took place in the first half of 2020 (see Roberts 2020a). It fails even more spectacularly for later waves (Roberts 2020b).

In this section we apply the logistic model to the four waves in South Africa. Solutions were found by numerical minimization of either the sum of the squares of the differences between model and data (least squares fit, or “LSQ”), or the sum of the absolute values of the differences (“L1”). These solutions were compared to plots of the seven day moving average of the data for each wave, and discordant solutions were discarded.^4^

### 2.4. RSA Wave 1

**Fig. 3.**
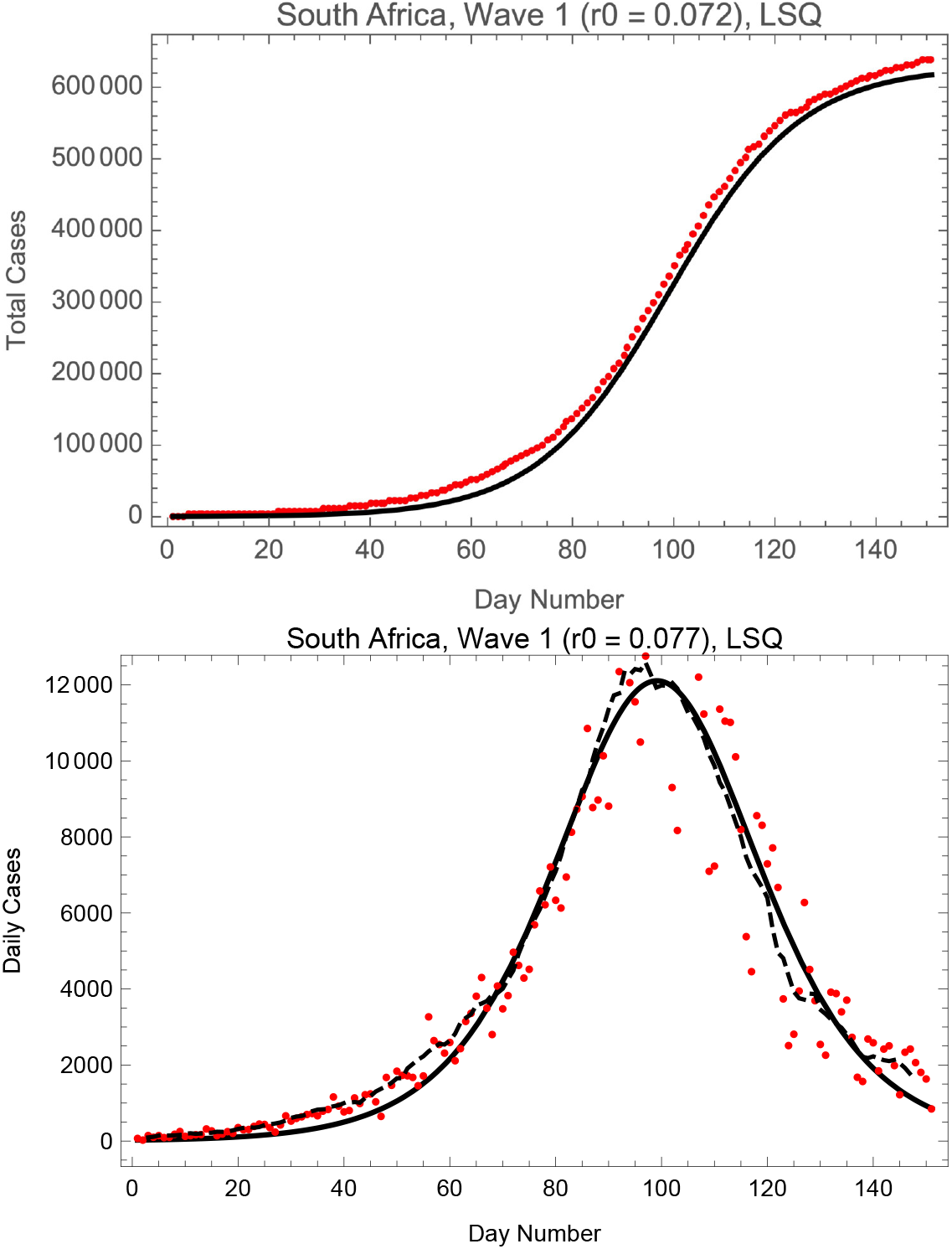
Fits of the logistic model to RSA cases of COVID-19 in wave 1. Day 1 is 11 April 2020. The broken line is the seven day moving average of the data.

### 2.5. RSA Wave 2

**Fig. 4.**
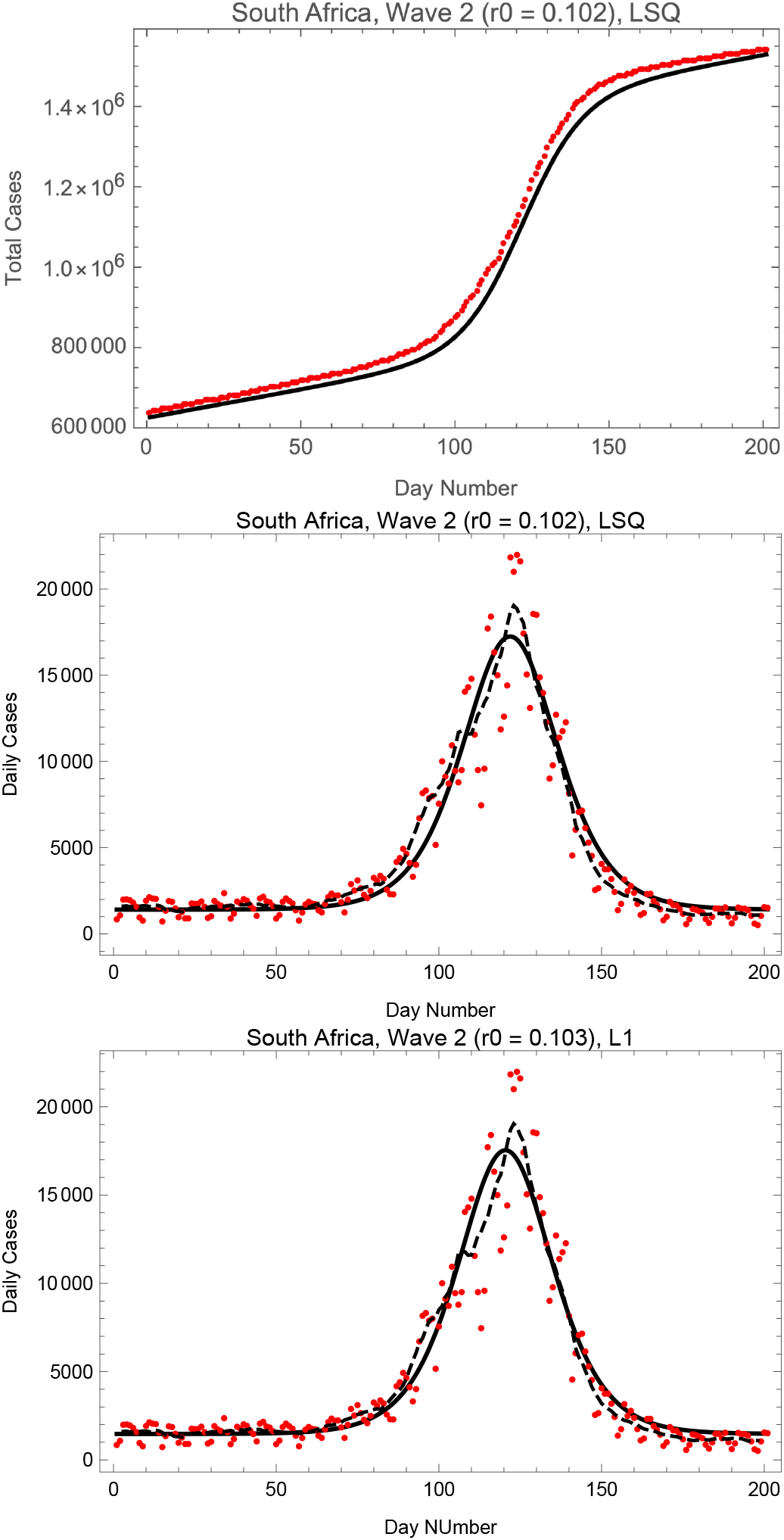
Fits of the logistic model to RSA cases of COVID-19 in wave 2. Day 1 is 8 September 2020. The broken line is the seven day moving average of the data.

### 2.6. RSA Wave 3

**Fig. 5.**
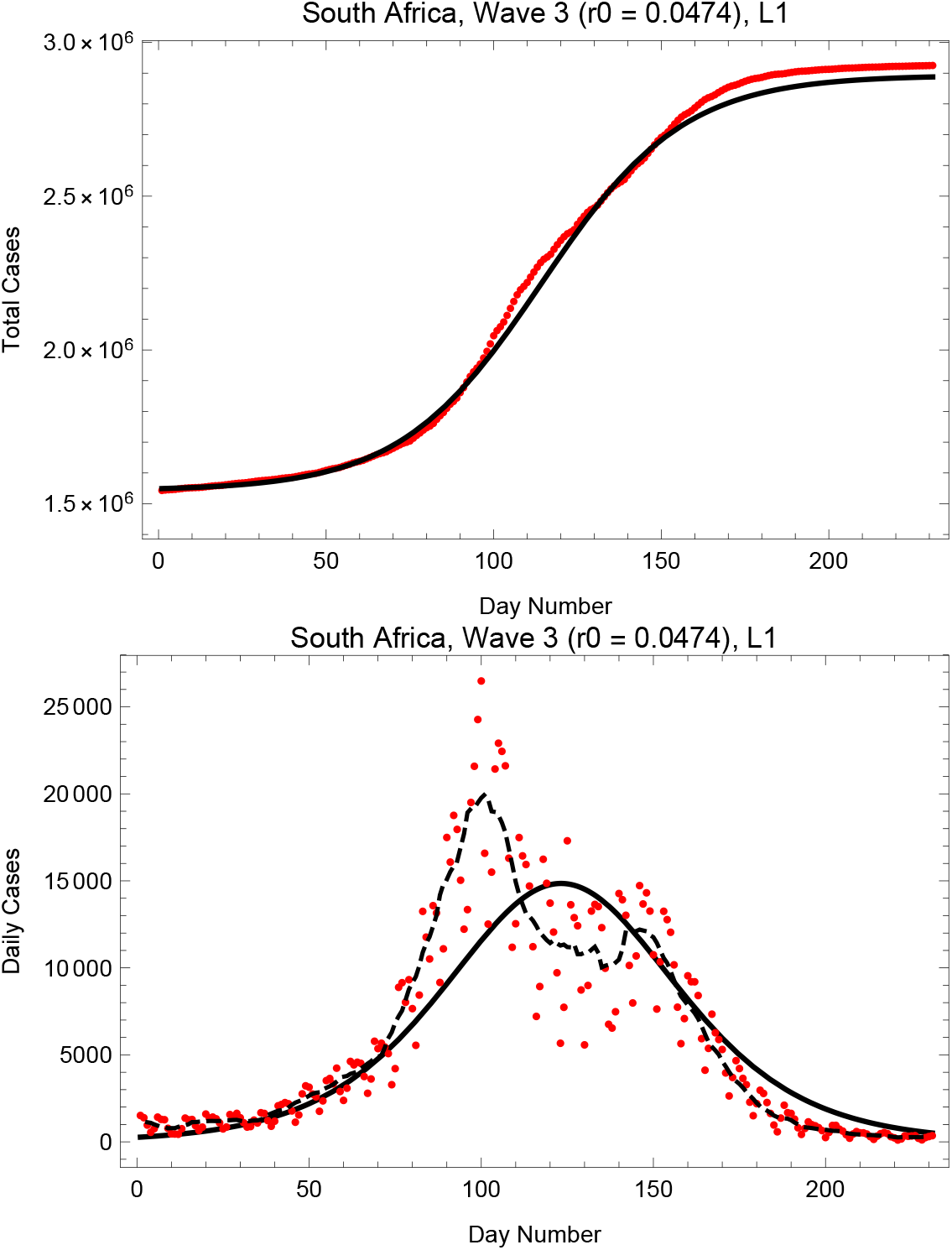
Fits of the logistic model to RSA cases of COVID-19 in wave 3. Day 1 is 12 November 2021. The broken line is the seven day moving average of the data.

### 2.7. RSA Wave 2

**Fig. 6.**
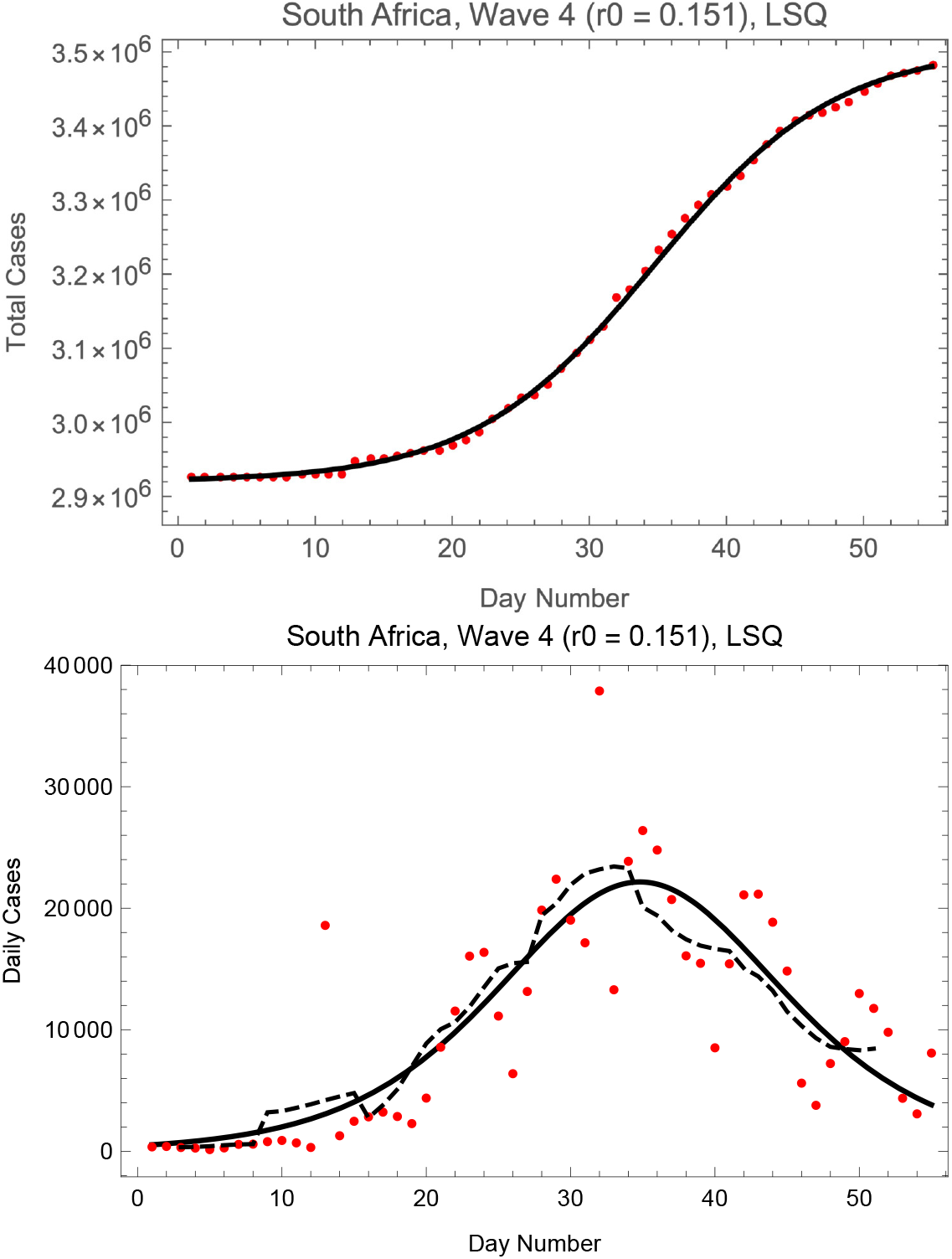
Fits of the logistic model to RSA cases of COVID-19 in wave 4. Day 1 is 12 November 2021. The broken line is the seven day moving average of the data.

## 3. Correlation of the Growth Rate and Duration for the COVID-19 Waves in the Republic of South Africa

It is natural that the growth rate and the duration of the waves of a pandemic should be correlated, higher growth rates leading to shorter durations. It can be shown (see the Appendix) that for a logistic model the daily cases distribution has a full-width-half-maximum given by

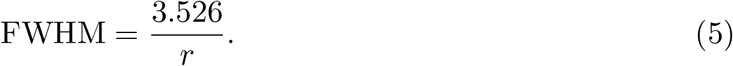

Note that this result is independent of *f*_0_ and *K*. The data and the prediction of Eq. 5 are compared in Fig. 7, where we see that the agreement is excellent. The full-width-half-maxima were estimated from curves of seven day moving averages of the daily data. The values of *r* for each wave were found by averaging those of the LSQ and L1 fits.

**Fig. 7.**
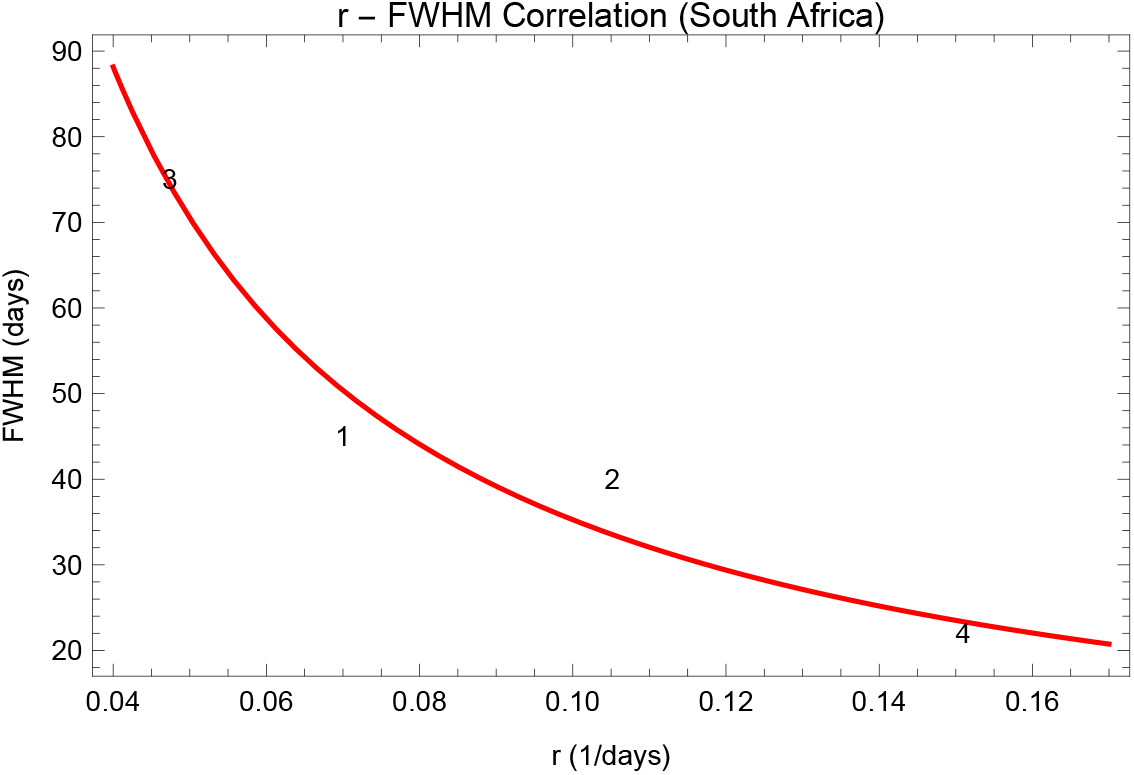
Correlation of the growth rate *r* and the FWHM of each wave for the COVID-19 pandemic in the Republic of South Africa. The curve is the relationship defined by Eq. 5.

## 4. Six Waves in the United States

In this section we apply the logistic model to five of the waves of COVID-19 in the United States, using the techniques described above. The fourth wave (around day 450 on Fig. 1) is omitted because we found it impossible to determine its full-width-half-maximum in a reliable way.

### 4.1. USA Wave 1

**Fig. 8.**
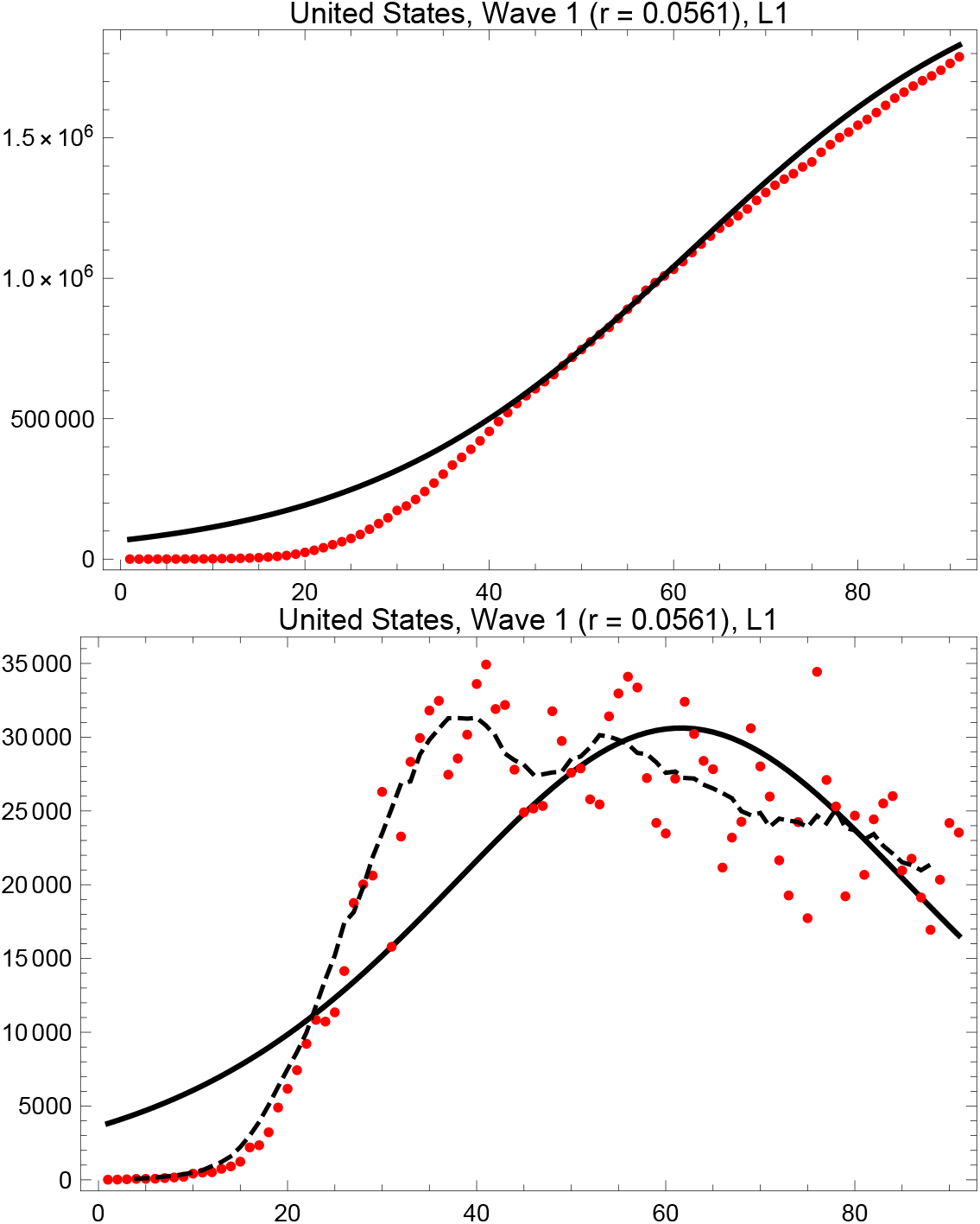
L1 fits of the logistic model to USA cases of COVID-19 in wave 1. Day 1 is 20 March 2020. The broken line is the seven day moving average of the data.

### 4.2. USA Wave 2

**Fig. 9.**
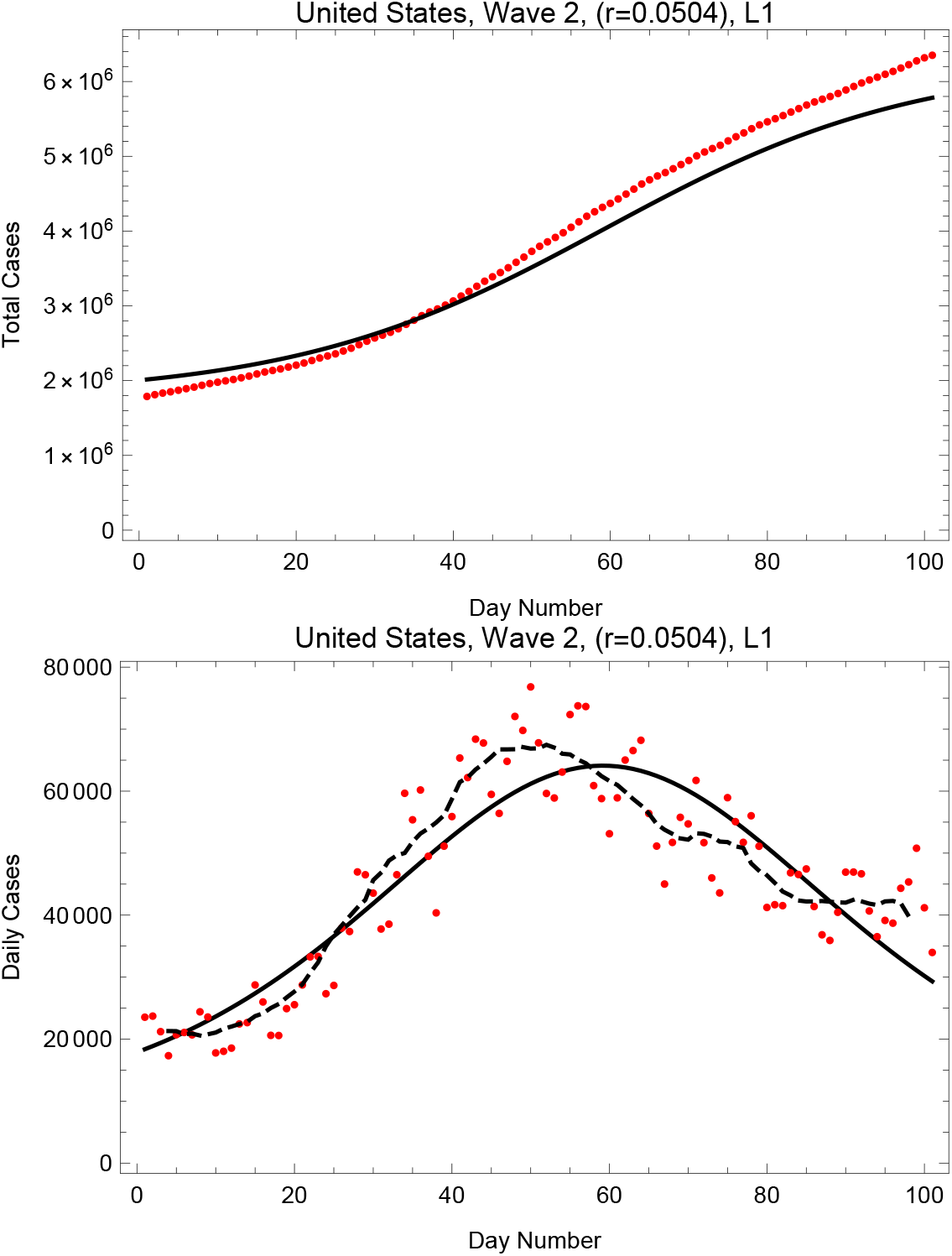
L1 fits of the logistic model to USA cases of COVID-19 in wave 2. Day 1 is 20 March 2020. The broken line is the seven day moving average of the data.

### 4.3. USA Wave 3

**Fig. 10.**
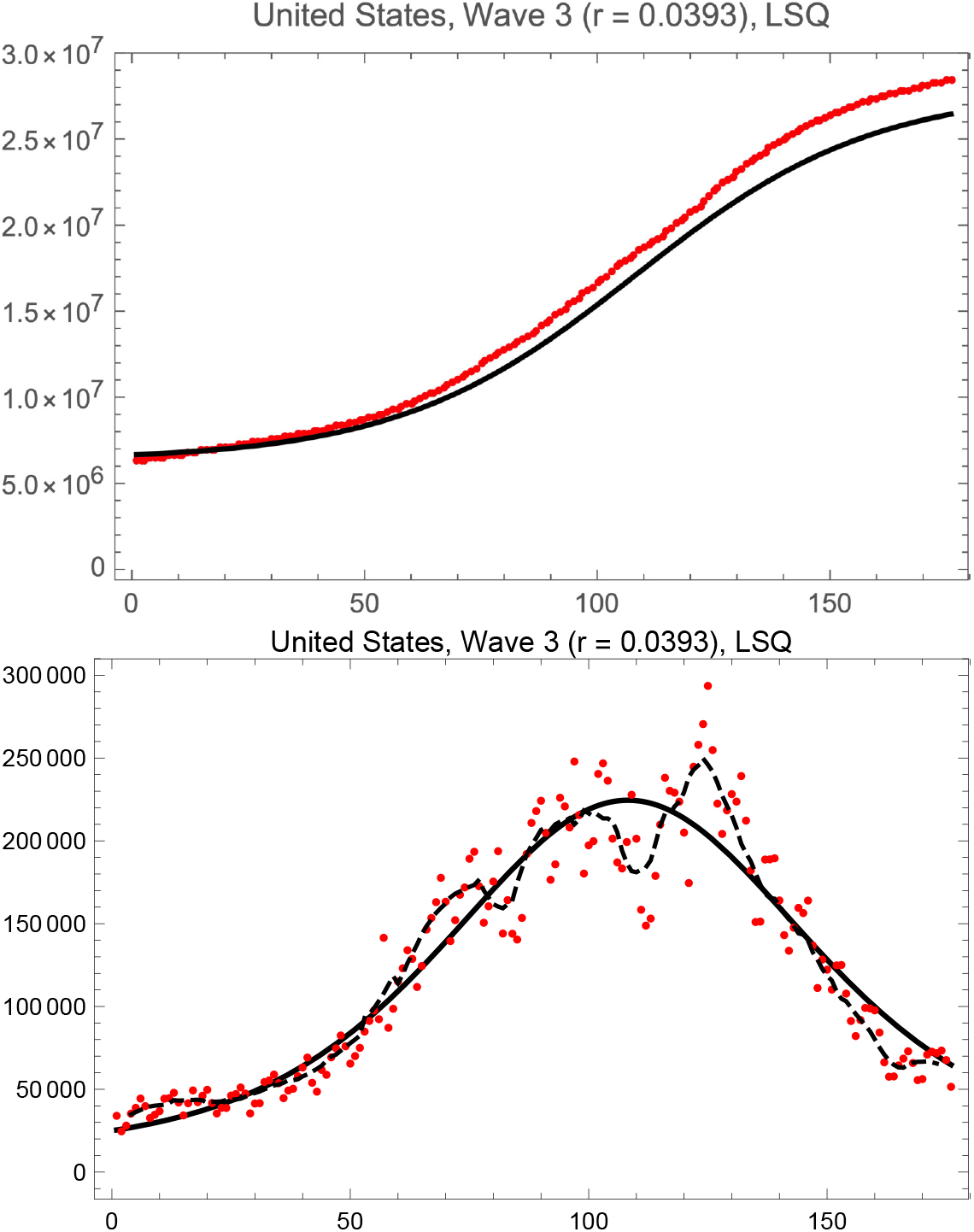
LSQ fits of the logistic model to total USA cases of COVID-19 in wave 3. Day 1 is 8 September 2020. The broken line is the seven day moving average of the data.

### 4.4. USA Wave 5

**Fig. 11.**
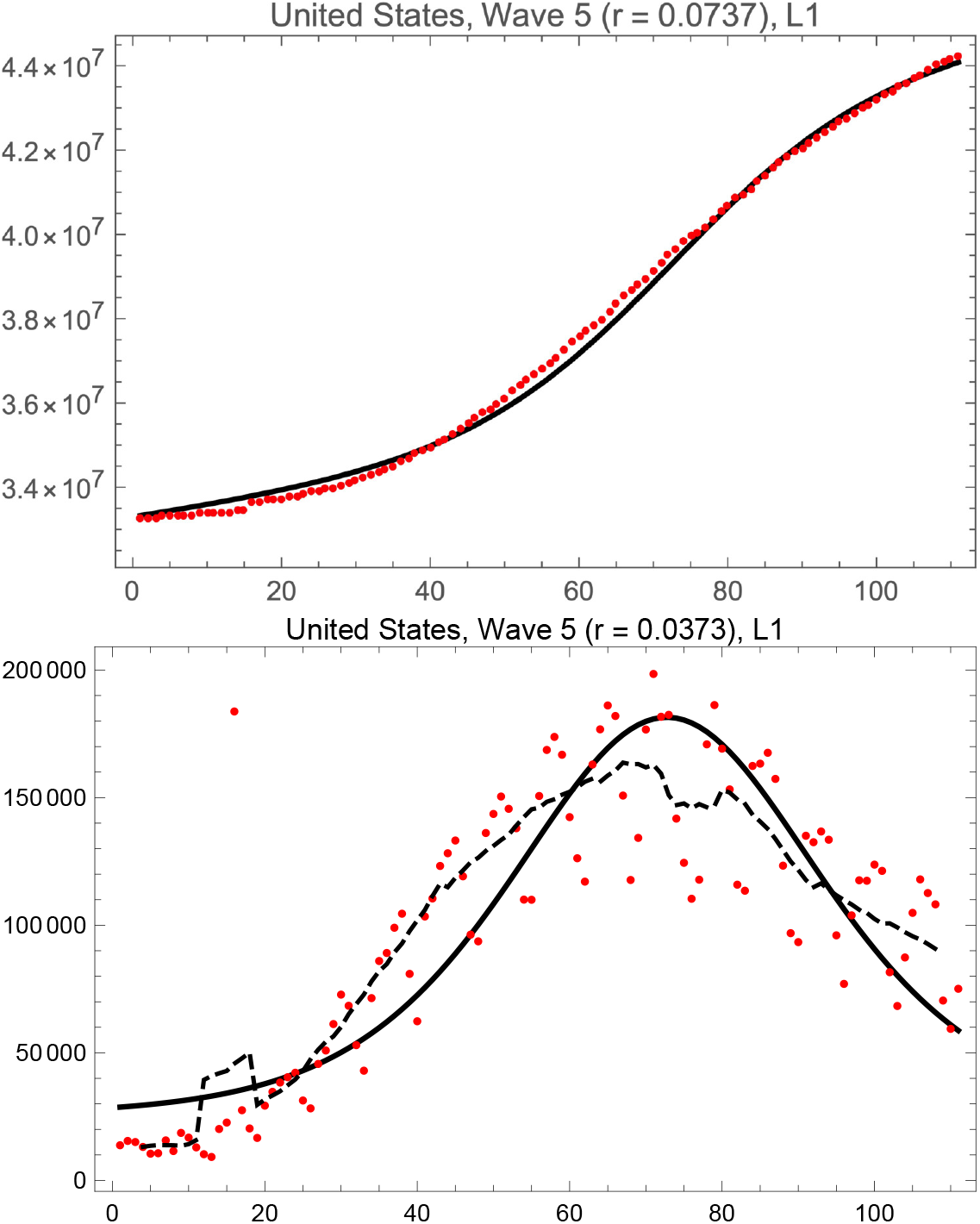
L1 fits of the logistic model to USA cases of COVID-19 in wave 5. Day 1 is 25 June 2021. The broken line is the seven day moving average of the data.

### 4.5. USA Wave 6

**Fig. 12.**
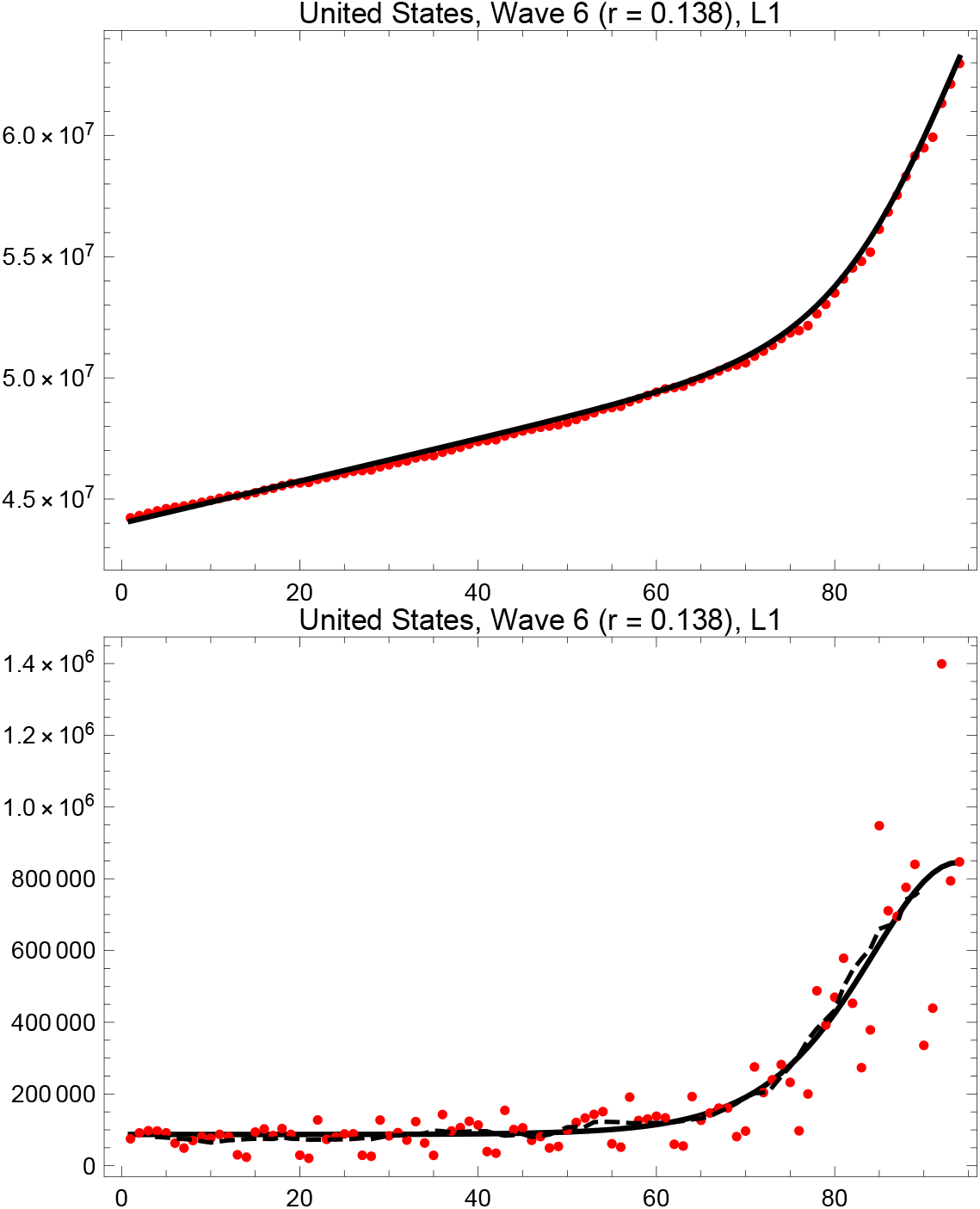
L1 fits of the logistic model to total USA cases of COVID-19 in wave 6. Day 1 is 13 October 2021. The broken line is the seven day moving average of the data.

**Fig. 13.**
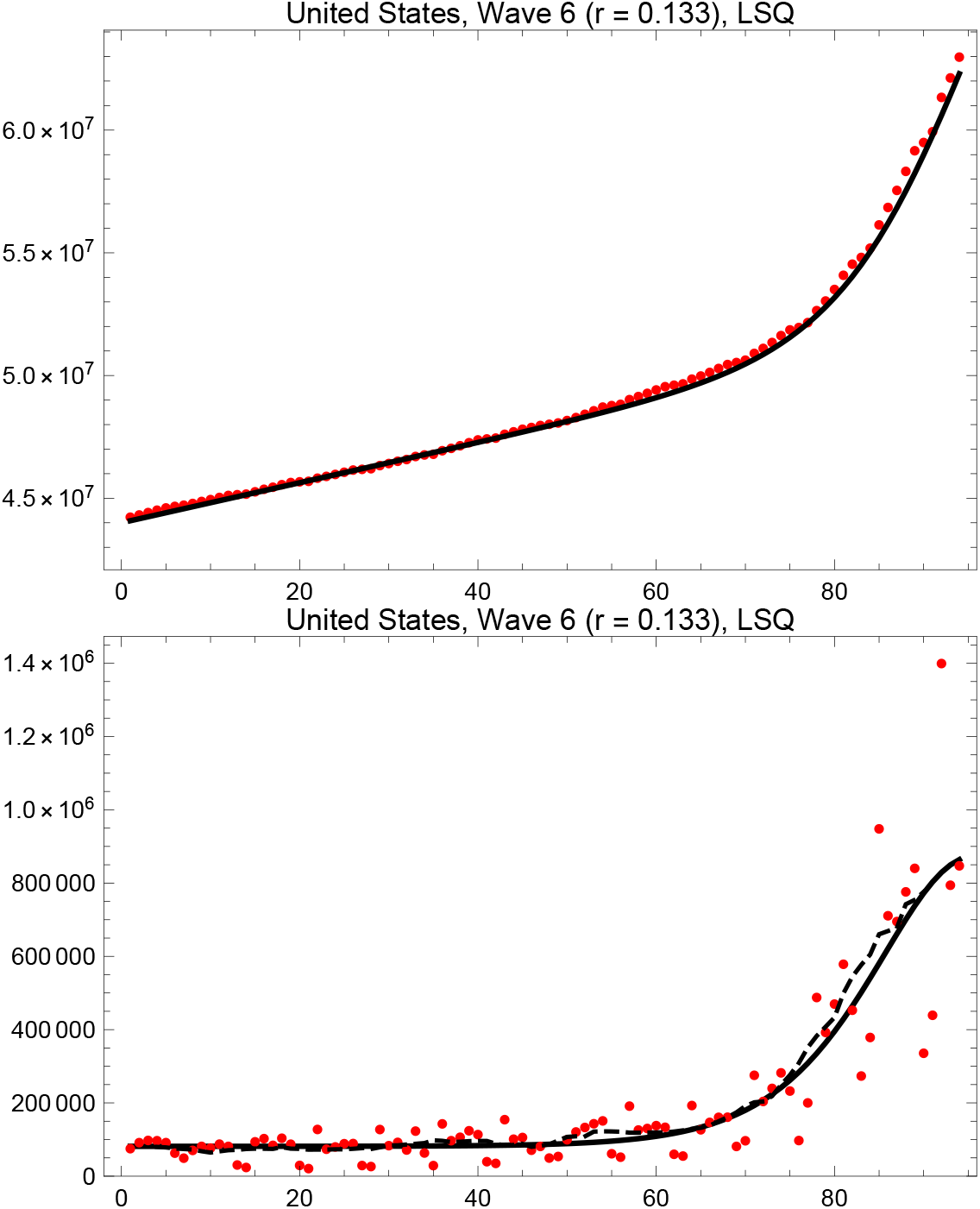
LSQ fits of the logistic model to USA cases of COVID-19 in wave 6. Day 1 is 13 October 2021. The broken line is the seven day moving average of the data.

## 5. Predictions for United States Wave 6

### 5.1. Comparison of Theory and Data

Fig. 14 shows the correlation between growth and duration of the various waves in the United States. The curve is the theoretical prediction of Eq. 5, identical to the one in Fig. 7. The agreement is not as good as for the South African data, but this so be expected because measurement of the FWHM of the data is very uncertain for two of the waves (examine Figs. 9 & 11).

**Fig. 14.**
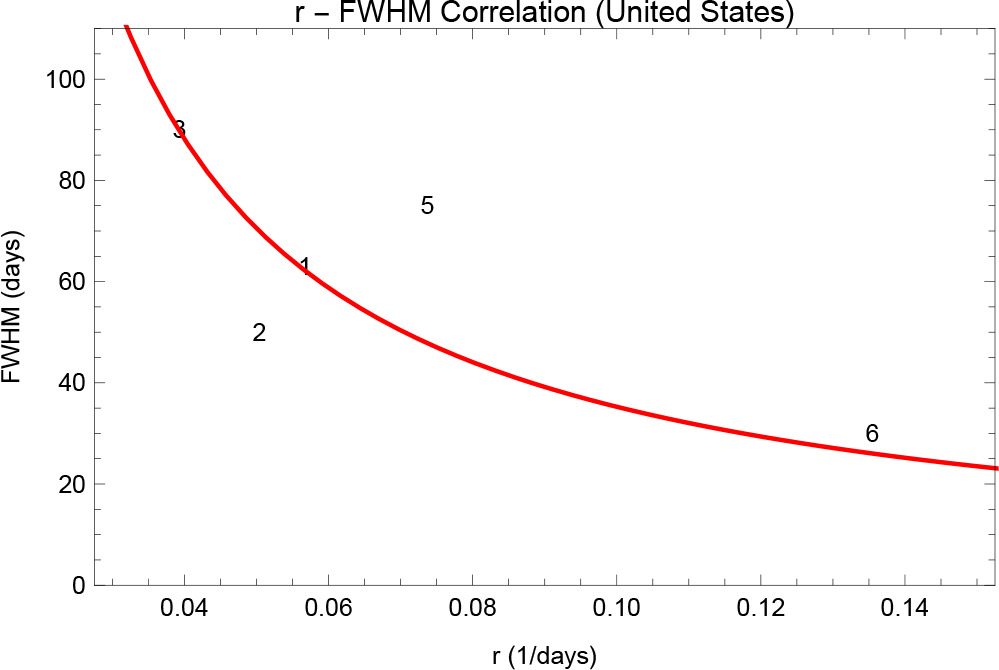
Correlation of the growth rate *r* and the FWHM of waves 1, 2, 3, 5 & 6 of the COVID-19 pandemic in the United states. The curve is the prediction of Eq. 5.

### 5.2. Prediction for the Omicron Wave in the United States

#### 5.2.1. Predictions from the Data

In Figs. 15 & 16 we show the predictions for wave 6 that follow from the L1 and LSQ solutions. The two provide roughly equivalent fits to the data (see Figs. 12 & 13), so we show the predictions made from each to exhibit some of the uncertainty. Examining the plot of the moving average of the wave 6 daily cases data we find a peak at day 95 and a FWHM of 25 days. From Fig. 15 we see that the total number of cases by the end of wave 6 is predicted to be 24 million.

We rather arbitrarily define the date at which the wave will be substantially over to be at one full-width-half-maximum past the peak, when the daily number of cases will be about 11% of its maximum. Thus using these data we expect that to be about day 120, which is 10 February 2022.

**Fig. 15.**
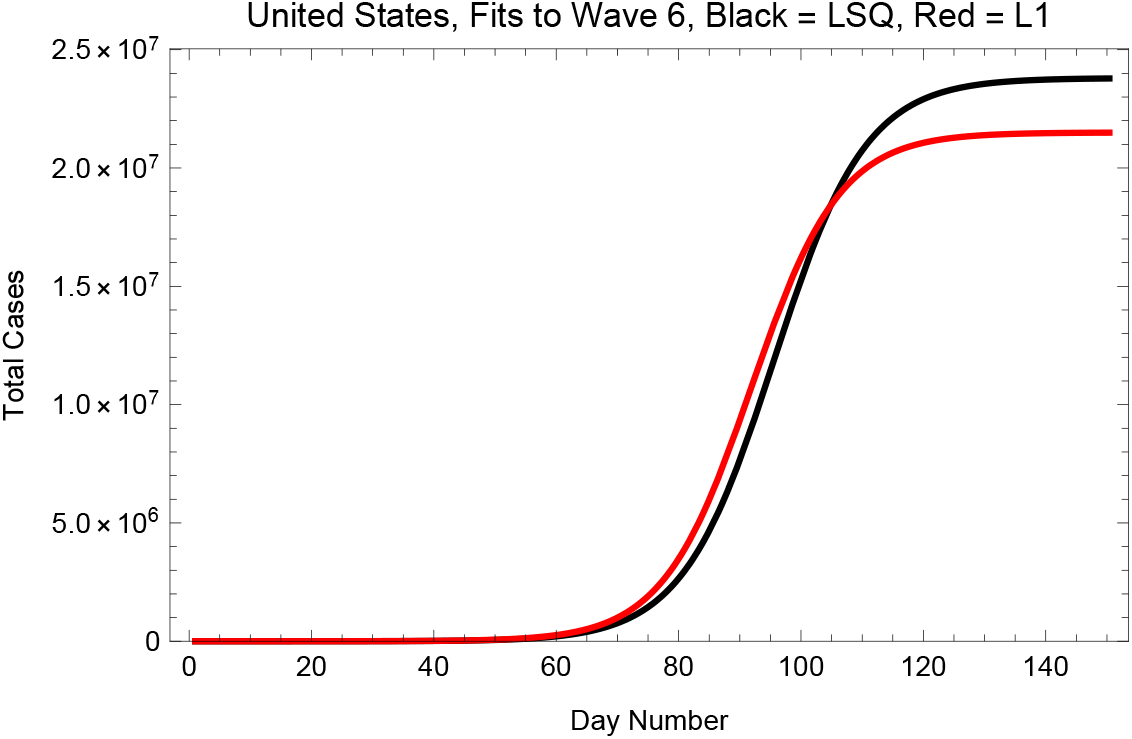
Predictions of the logistic model for the total USA cases of COVID-19 in wave 6. The final total numbers of cases are 23.8 million for the LSQ fit and 21.5 million for the L1 fit.

**Fig. 16.**
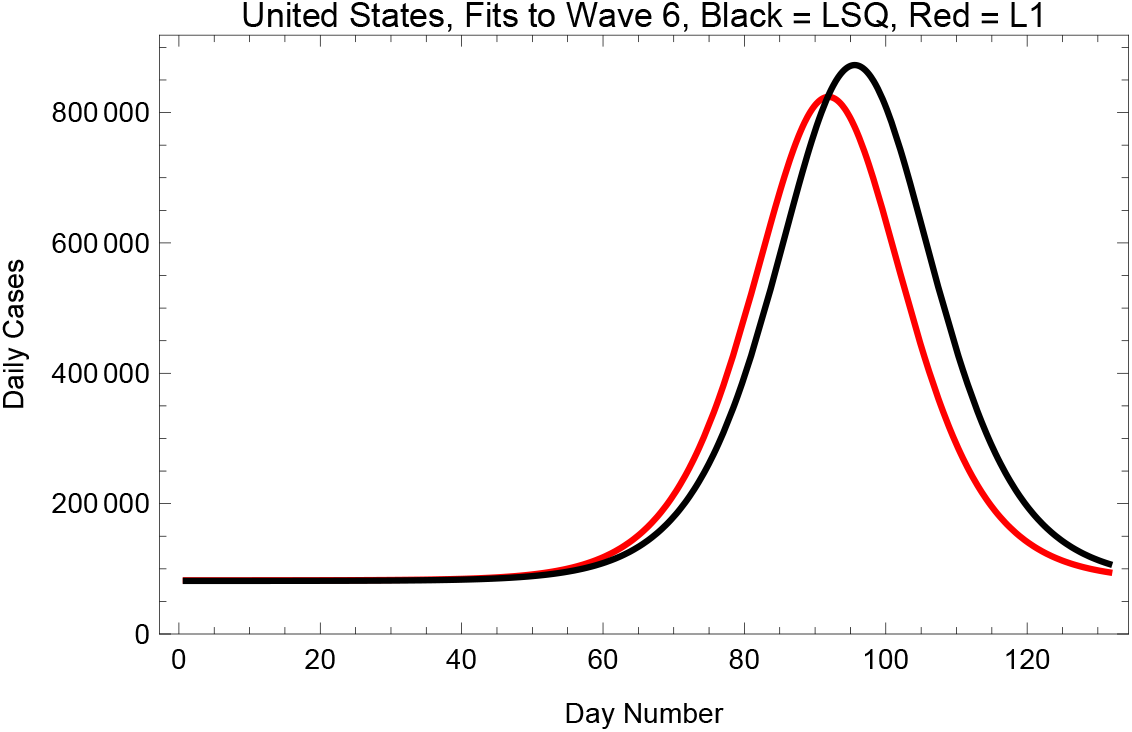
Predictions of the logistic model for the daily USA cases of COVID-19 in wave 6. The peaks are at days 95.6 for LSQ fit and 91.9 for the L1 fit.

#### 5.2.2. Predictions from the Theory

We can use Eqs. 8 & 10 and the values of *f*_0_, *r*, and *K* to find the expected date of the peak of wave 6 and its FWHM. Adopting the LSQ fit to this wave (*f*_0_= 71.5 days, *K* = 2.38 × 10^7^, *r* = 0.133 days^−1^) this leads to an expected day of peak daily cases at 95.6 days and an expected FWHM of 26.5 days, and thus a date of substantial diminution of day 122 (12 February 2022). For the L1 solution (*f*_0_= 51.5 days, *K* = 2.20 × 10^7^, *r* = 0.138 days^−1^) these are 93.9 days and 25.6 days, leading to a date of day 120 (10 February 2022). These are in good accord with the estimates obtained above. Thus this approach suggests that wave 6 will be substantially over by day 121, or 11 February 2022.

## 6. Conclusions

In this paper we have used the logistic model for epidemics to describe the COVID-19 outbreaks in the Republic of South Africa and in the United States. We find a universal analytic relationship between the growth rates and the durations of each wave, and this is closely followed by the (very clean) data from South Africa. When applied to the United States, the data are messier, but a tentative prediction is possible for the expected duration of the current Omicron wave in the US – it should be substantially over by about day 121, or about 11 February 2022. By the full end of the Omicron wave the total number of infected persons in the US is projected to be between 22 and 24 million.

## Data Availability

WHO COVID-19 Website

https://covid19.who.int/WHO-COVID-19-global-data.csv

## 7. Acknowledgements

We thank Brian Boyle, Mary Roberts, and Bob Sauer for their helpful comments and suggestions.

## 9. Appendix Derivation of Equation 4

The differential distribution for the logistic function is^5^

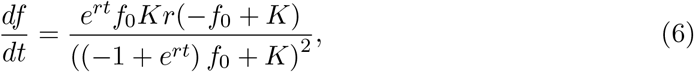

and is derivative is

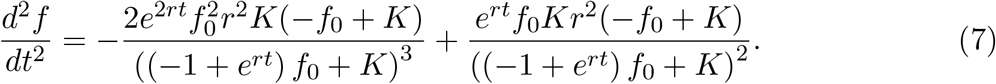

This is zero at

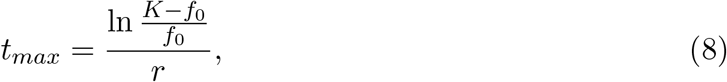

and *df* / *dt* at this maximum is

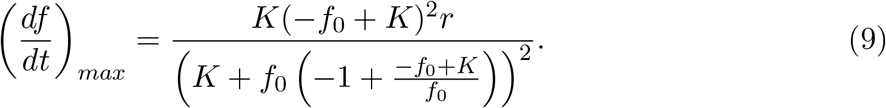

Setting *df* / *dt* equal to half of this value and solving for the locations yields a complicated expression that reduces to

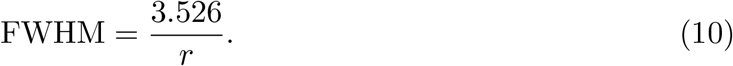

Note that neither *f*_0_ nor *K* enters this result.

## Footnotes

2 This is a version of the Bernoulli differential equation *f* ^*t*^ = *f* (1 − *f*).

3 Fitting the daily numbers is statistically the preferred procedure as the data points are independent, unlike those for daily totals.

4 In a numerical minimization of a non-linear function of multiple variables there is always a possibility of erroneous solutions.

5 All the calculations were done with Mathematica.

